# Greater baseline cortical atrophy in the dorsal attention network predicts faster clinical decline in Posterior Cortical Atrophy

**DOI:** 10.1101/2024.10.15.24315270

**Authors:** Yuta Katsumi, Ryan Eckbo, Marianne Chapleau, Bonnie Wong, Scott M. McGinnis, Alexandra Touroutoglou, Bradford C. Dickerson, Deepti Putcha

## Abstract

**Background and Objectives:** Posterior Cortical Atrophy (PCA) is a clinical syndrome characterized by progressive visuospatial and visuoperceptual impairment. As the neurodegenerative disease progresses, patients lose independent functioning due to the worsening of initial symptoms and development of symptoms in other cognitive domains. The timeline of clinical progression is variable across patients, and the field currently lacks robust methods for prognostication. Here, evaluated the utility of MRI-based cortical atrophy as a predictor of longitudinal clinical decline in a sample of PCA patients.

**Methods:** PCA patients were recruited through the Massachusetts General Hospital Frontotemporal Disorders Unit PCA Program. All patients had cortical thickness estimates from baseline MRI scans, which were used to predict longitudinal change in clinical impairment assessed by the CDR Sum-of-Boxes (CDR-SB) score. Multivariable linear regression was used to estimate the magnitude of cortical atrophy in PCA patients relative to a group of amyloid-negative cognitively unimpaired participants. Linear mixed-effects models were used to test hypotheses about the utility of baseline cortical atrophy for predicting longitudinal clinical decline.

**Results:** Data acquired from 34 PCA patients (mean age = 65.41 ± 7.90, 71% females) and 24 controls (mean age = 67.34 ± 4.93, 50% females) were analyzed. Sixty-two percent of the PCA patients were classified as having mild cognitive impairment (CDR 0.5) at baseline, with the rest having mild dementia (CDR 1). Each patient had at least one clinical follow-up, with the mean duration of 2.78 ± 1.62 years. Relative to controls, PCA patients showed prominent baseline atrophy in the posterior cortical regions, with the largest effect size observed in the visual network of the cerebral cortex. Cortical atrophy localized to the dorsal attention network, which supports higher-order visuospatial function, selectively predicted the rate of subsequent clinical decline.

**Discussion:** These results demonstrate the utility of a snapshot measure of cortical atrophy of the dorsal attention network for predicting the rate of subsequent clinical decline in PCA. If replicated, this topographically-specific MRI-based biomarker could be useful as a clinical prognostication tool that facilitates personalized care planning.

## Introduction

Posterior Cortical Atrophy (PCA) is a clinical syndrome characterized by progressive impairment in visuoperceptual, spatial, and other cognitive functions with key localization in occipito-parietal and/or occipito-temporal regions^1^. PCA typically presents in the mid-50s to mid-60s, with a heterogeneous symptom constellation including visual and/or spatial processing impairments, dyscalculia, dyslexia, dysgraphia, apraxia, and/or agnosia for objects or faces^2,3^. In the vast majority of PCA cases, these symptoms arise as a result of Alzheimer’s disease (AD) pathologic changes (∼94%)^2^. Thus, PCA syndrome has been referred to as the “visual variant” of AD^4^. Although cognitive domains outside of visual functioning are thought to be spared at the earliest stages of the illness, symptoms in other domains including episodic memory, executive functions, and language eventually become evident and often have a significant impact on daily functioning as the disease progresses. The timeline of this progression over time is highly variable across patients, and the field currently lacks robust methods for prognostication. In this new era of disease-modifying therapies for AD, there is an urgent need to develop accurate methods for prognostication and outcomes monitoring to inform clinical trial design as well as individualized treatment recommendations appropriate for PCA syndrome. To the best of our knowledge, very few studies have investigated factors predicting the trajectory of clinical (cognitive and functional) decline specifically in PCA. Compared with typical amnestic AD, PCA patients exhibited more severe visuospatial and visuoperceptual impairments at baseline and also declined faster in these domains over time^5,6^. Poorer baseline performance on the Montreal Cognitive Assessment Battery (MoCA)^7^ in PCA was associated with faster longitudinal decline in visuospatial performance, suggesting that those with more severe global cognitive impairment at baseline subsequently declined at a faster rate within the visuospatial domain^8^.

Prior work from our group and others investigating a variety of neurodegenerative syndromes has demonstrated the utility of baseline imaging features for predicting the rate of subsequent clinical decline^8–13^. In a series of recent studies, we highlighted the clinical prognostic value of neurodegenerative and neuropathologic changes in specific functional networks of the cerebral cortex^14,15^. First, in a sample of patients with early-stage Primary Progressive Aphasia (PPA), we demonstrated that the magnitude of cortical atrophy in the language network and the frontoparietal network predicted clinical progression from the mild cognitive impairment (MCI) to the dementia stage of PPA^14^. While prominent left-lateralized atrophy affecting the perisylvian regions of the language network at baseline also predicted subsequent clinical progression, we found that the strongest predictor of progression to dementia was atrophy in the left frontoparietal network, which was less atrophic at baseline compared with the language network. In another study of patients with early-stage AD presenting with a variety of atypical clinical syndromes including PCA, we showed that the accumulation of neurofibrillary tangles composed of aggregated hyperphosphorylated tau in the default mode network strongly predicted clinical decline one year later^15^. While abnormal tau accumulation at baseline was primarily localized to the posterior regions of the default mode network, faster clinical decline was associated with greater tau burden in the broader default mode network, including its anterior nodes in the medial and lateral prefrontal cortex. Taken together, these findings suggest that baseline cortical atrophy and tau deposition in specific cortical functional networks could be useful as early prognostic biomarkers of subsequent clinical decline. Some studies of atypical AD including PCA have identified the contribution of baseline neuroimaging measures to predicting future clinical impairment over and above that of baseline *clinical* measures^10,15^, suggesting that the incorporation of imaging features improves the accuracy of clinical prognostication.

PCA due to AD is typically characterized by prominent atrophy localized in the occipital and posterior temporo-parietal cortical areas that are components of multiple functional networks, including the visual network as well as the posterior nodes of the dorsal attention network (DAN), default mode network, and frontoparietal network^3,16,17^. As the disease progresses, AD-related accumulation of tau pathology spreads anteriorly to the frontal cortex (e.g., frontal eye fields)^18–20^. Anterior progression in the topography of neurodegeneration over time, characterized by greater involvement of lateral temporo-parietal and medial parietal areas^5,19,20^, has also been described in PCA. Taken together, these findings suggest that, as the visual network becomes saturated with AD-related neuropathologic and neurodegenerative changes early in the disease course, these changes continue to spread to dorsal and anterior regions within the DAN, default mode network, and frontoparietal network. The specific topography of cortical atrophy that is predictive of future clinical decline in PCA, however, remains unclear.

In the present study, we sought to investigate the prognostic utility of baseline cortical atrophy for predicting the rate of longitudinal clinical (cognitive and functional) decline in a sample of patients with PCA at early symptomatic stages of illness. On the basis of evidence reviewed above, we hypothesized that PCA patients would exhibit prominent atrophy at baseline in posterior cortical regions including those comprising the visual network and posterior components of the DAN, default mode network, and frontoparietal network. Building upon prior work pointing to the anterior progression of tau pathology and neurodegeneration in PCA, we further hypothesized that baseline atrophy in the temporo-parietal regions that are part of the DAN, default mode network, and frontoparietal network, but *not* the visual network, would be predictive of longitudinal clinical decline. If supported, this work would highlight the prognostic power of regional baseline cortical atrophy measures in early-stage PCA due to AD, laying the groundwork for the use of this measure in patient-centered discussions of prognosis and treatment planning.

## Methods

### Participants

The present study included 34 individuals who fulfilled diagnostic criteria for PCA^3,21,22^, all of whom were recruited from the Massachusetts General Hospital (MGH) Frontotemporal Disorders Unit PCA program^23,24^. Of the 34 PCA patients, 29 had biomarker or pathological evidence consistent with underlying AD pathology as determined by amyloid-PET (*n* = 21), cerebrospinal fluid (CSF) (*n* = 3), or autopsy-proven ADNC (*n* = 5). The molecular biomarker status of the remaining five individuals is unknown, with a high degree of suspicion for ADNC based on clinical and neuroimaging profiles. All patients received a comprehensive clinical evaluation comprising a structured history obtained from both patient and informant to inform clinician scoring on the Clinical Dementia Rating (CDR) scale^25^. They also underwent a comprehensive neurological and psychiatric examinations and neuropsychological assessment. Clinical diagnostic formulation was performed through consensus conference by our multidisciplinary team of neurologists, neuropsychologists, and speech and language pathologists, with each patient classified based on all available clinical information as having a 3-step diagnostic formulation of MCI or dementia (Cognitive Functional Status), a specific Cognitive-Behavioral Syndrome, and a likely etiologic neuropathologic diagnosis^26,27^.

We additionally included a group of 24 amyloid-negative (Aβ-) cognitively unimpaired (CU) control participants, all of whom had normal brain structure based on MRI and low cerebral amyloid based on quantitative analysis of PiB PET data (FLR DVR < 1.2). This control sample was used as a reference for quantifying the magnitude of baseline cortical atrophy in PCA patients. Individuals were excluded from our patient and control groups if they had a primary psychiatric or other neurologic disorder including major cerebrovascular infarct or stroke, seizure, brain tumor, hydrocephalus, multiple sclerosis, HIV-associated cognitive impairment, or acute encephalopathy.

### Assessment of clinical impairment

Each PCA patient’s level of clinical impairment was assessed by the CDR Sum-of-Boxes (CDR-SB) score. On average, baseline CDR-SB scores were obtained from each patient within 0.69 ±1.04 months relative to the date of MRI. Each patient had at least one follow-up, with the mean follow-up duration of 2.78 ± 1.62 years.

### MRI data acquisition and preprocessing

At baseline, brain structural MRI data were acquired from each participant on a Siemens 3 Tesla scanner (either Prisma Fit or Tim Trio) using a T1-weighted magnetization prepared rapid acquisition sequence (MPRAGE) (repetition time [TR] = 2300 ms, echo time [TE] = 2.98 ms, flip angle = 9°, slice thickness = 1 mm, field of view [FOV] = 240 × 256 mm^2^) or a multi-echo MPRAGE sequence (TR = 2530 ms, TEs = 1.64/3.5/5.36/7.22 ms, flip angle = 7°, slice thickness = 1 mm, FOV = 256 × 256 mm^2^). All MRI data were visually inspected for gross artifacts (e.g., head motion) and evaluated on image quality prior to data processing. After passing quality control, each participant’s (ME)MPRAGE data underwent intensity normalization, skull stripping, and an automated segmentation of cerebral white matter to locate the gray matter/white matter boundary via FreeSurfer v6.0 (https://surfer.nmr.mgh.harvard.edu). Defects in surface topology were corrected and the gray/white boundary was deformed outward using an algorithm designed to obtain an explicit representation of the pial surface. We visually inspected each participant’s cortical surface reconstruction for technical accuracy. Estimates of cortical thickness were calculated as the closest distance from the gray/white boundary to the gray/CSF boundary at each vertex on the tessellated surface^28^. Individual maps of cortical thickness were registered to template surface space (*fsaverage*) and smoothed geodesically with a full-width half-maximum (FWHM) of 10 mm.

### Statistical analysis

We conducted all statistical analyses of demographic, clinical, and neuroimaging data using *R* version 4.2.1. We compared demographic variables between PCA patients and Aβ-CU participants using two-tailed independent samples *t*-tests for age and education and a chi-square test for sex. Group differences were considered statistically significant if *p* < .05. Measures of effect size were expressed as Cohen’s *d* (*t*-tests) and Cohen’s *w* (chi-square tests). To estimate the rate of longitudinal change in CDR-SB scores in PCA patients, we constructed a linear mixed-effects model using the *lmer* function from the *lme4* package (version 1.1-35.1)^29^. This model was fit by maximizing the restricted likelihood and included time since baseline (in years) as the only fixed predictor with random intercepts and slopes for participants. Additionally, we constructed a mixed-effects model with the interaction term between time since baseline and baseline CDR-SB scores (Time*CDR-SB_baseline_) to examine the relationship between baseline CDR-SB scores and the rate of change in this measure over time.

Using individual cortical thickness maps as inputs, we created whole-cortex vertex-wise multivariable linear regression models to identify areas of the cerebral cortex where PCA patients showed abnormal cortical thickness (i.e., atrophy) relative to Aβ-CU participants at baseline, while controlling for baseline age and sex as covariates of no interest. Next, to examine the prognostic utility of baseline cortical atrophy for predicting longitudinal clinical decline, we constructed vertex-wise linear mixed-effects models. These models were fit by maximizing the restricted likelihood and included the interaction term between time since baseline (follow-up duration) and baseline cortical thickness (Time*Thickness), baseline age and sex as covariates of no interest, and random intercepts and slopes for participants. For all vertex-wise linear models, statistical significance was assessed at a vertex-wise threshold of *p* < .05 corrected for multiple comparisons by controlling the false discovery rate (FDR).

To characterize the spatial topography of baseline cortical atrophy and its clinical prognostic utility in terms of large-scale functional networks of the cerebral cortex, we used an established cortical parcellation derived from a sample of 1,000 healthy young adults^30^. Based on the 7-network solution of this parcellation, we defined the following networks bilaterally: Visual, somatomotor, dorsal attention, ventral attention, limbic, frontoparietal, and default. Here, we adhere to the original and conventional use of the label, “limbic” network, although we recognize that cortical regions part of this network are often considered part of the default mode network^31–33^, and that both networks contain agranular, limbic tissue^34,35^. Using the spherical registration of each participant to *fsaverage* space, the seven network parcellations were registered back to individual participants’ native surface space. For each participant, mean cortical thickness was calculated within each network by averaging thickness estimates across all vertices within its boundaries.

Finally, we performed additional network-based characterization of our linear regression and mixed-effects model results by projecting the associated vertex-wise statistical maps to a flat map representation of *fsaverage* surface. We focused on the visual network and the DAN in this analysis given that the results of our statistical models highlight opposite patterns of involvement between these two networks. First, unthresholded vertex-wise maps of effect size estimates (i.e., a Cohen’s *d* map from comparing baseline thickness between PCA patients and Aβ-CU participants and a ΔSlope map resulting from a Time*Thickness interaction in PCA patients) were converted to *Z* scores. Using these maps, we extracted vertex-wise effect size estimates from the visual network (number of vertices = 44,151) and DAN (number of vertices = 35,115). Permutation tests were used to compare the mean effect sizes across networks within each map and between maps, with 10,000 random permutations of network labels per comparison.

### Standard Protocol Approvals, Registrations, and Patient Consents

The study design and protocol were approved by the Mass General Brigham Institutional Review Boards for human research. Each participant and their informant gave written informed consent in accordance with the Mass General Brigham Human Subjects Research Committee guidelines.

## Data Availability

Data not provided in the article because of space limitations may be shared (anonymized) at the request of any qualified investigator for purposes of replicating procedures and results.

## Results

### Demographic and clinical characteristics of the sample

Table 1. summarizes the demographic and clinical characteristics of the current sample. PCA patients and Aβ-CU participants did not significantly differ in age at baseline (*t* = 1.06, *p ≤* .29, *d* = 0.28), sex (*χ*^2^ (1, *N* = 58) = 1.73, *p ≤* .19, *w* = 0.21), or years of education (*t* = -1.79, *p ≤* .08, *d* = 0.59). Regarding cognitive functional status, the majority of the PCA patients included in this study were classified as having MCI (CDR 0.5) at baseline. The annualized rate of change in CDR-SB scores was estimated to be 2.28 points in the entire sample of PCA patients (range = [0.76, 4.73], 95% confidence interval = [1.81, 2.74], *p* < .0001). When stratified for baseline CDR global scores, the rate of change in CDR-SB scores was 1.9 points for PCA patients with CDR 0.5 (95% CI = [1.45, 2.35], *p* < .0001) and 2.78 points for those with CDR 1 (95% CI = [1.58, 3.99], *p* < .0001). Consistent with this result, the rate of longitudinal change in CDR-SB scores was associated with baseline scores at a trend level (*F*(1,30.733) = 3.00, *p ≤* .093), suggesting that those PCA patients with more severe clinical impairment at baseline tended to progress at a faster rate (**Figure 1**).

**Table 1.**
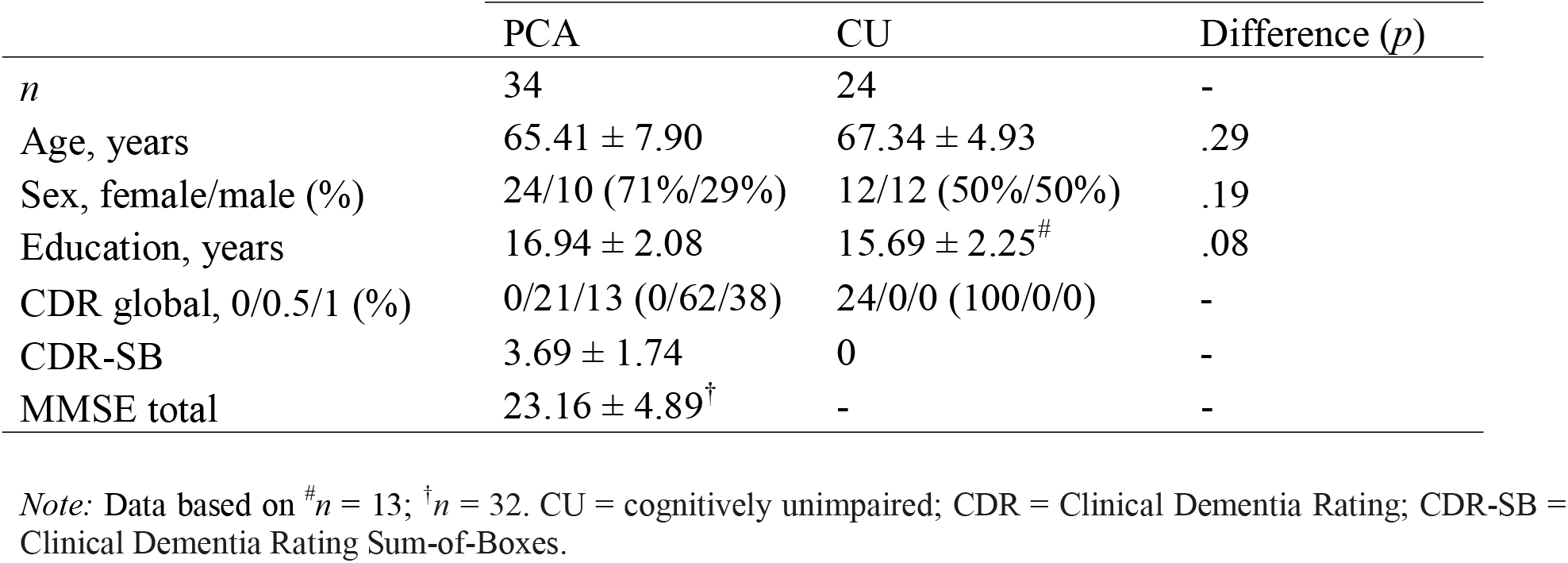
Sample characteristics at baseline.

**Figure 1.**
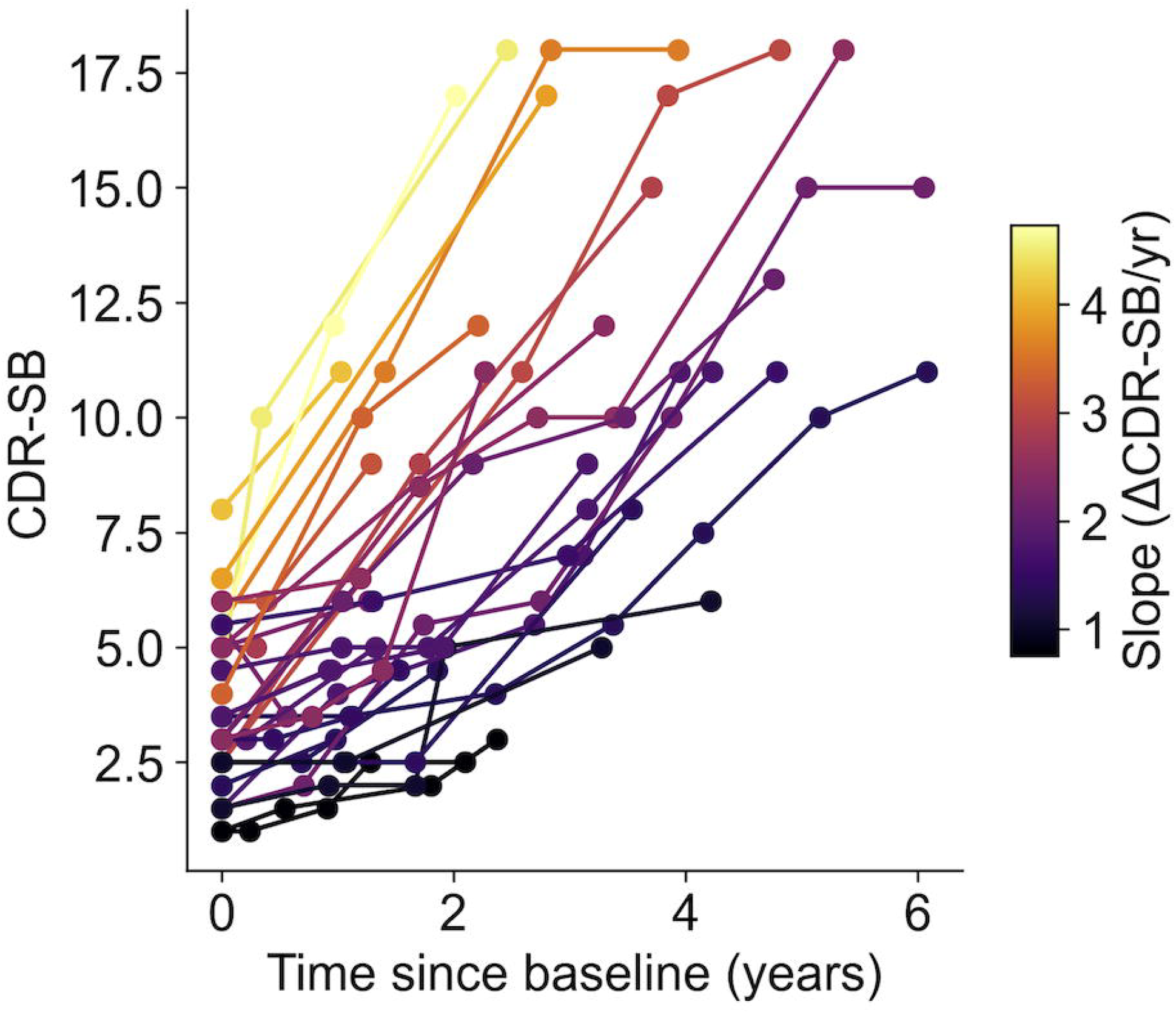
Longitudinal clinical decline in PCA patients. The spaghetti plot identifies CDR-SB scores across annual timepoints, where each line represents a PCA patient. Each line is color coded by the slope of longitudinal change in CDR-SB scores estimated from a linear mixed-effects model, with brighter colors indicating steeper decline.

### Spatial topography of baseline cortical atrophy

Relative to Aβ-CU participants, PCA patients showed prominent atrophy in the posterior cortical regions including the occipital, ventral and lateral temporal, and lateral and medial parietal cortex bilaterally. Modest atrophy was identified in the lateral frontal cortex bilaterally, uniquely localized to the frontal eye fields and inferior frontal junction (**Figure 2A**). At the overall network level, PCA patients exhibited the most prominent atrophy in the visual network (difference in thickness [Δ] = 0.31, *p* < .0001, *d* = 2.31) and DAN (Δ = 0.27, *p* < .0001, *d* = 1.87), followed by moderate atrophy in the frontoparietal (Δ = 0.11, *p ≤* .0004, *d* = 0.99), default mode (Δ = 0.096, *p ≤* .002, *d* = 0.86), ventral attention (Δ = 0.084, *p ≤* .003, *d* = 0.83), and limbic (Δ = 0.070, *p ≤* .042, *d* = 0.55) networks. We found no significant difference in cortical thickness between PCA patients and Aβ-CU participants in the somatomotor network (Δ = 0.030, *p ≤* .37, *d* = 0.24) (**Figure 2B**).

**Figure 2.**
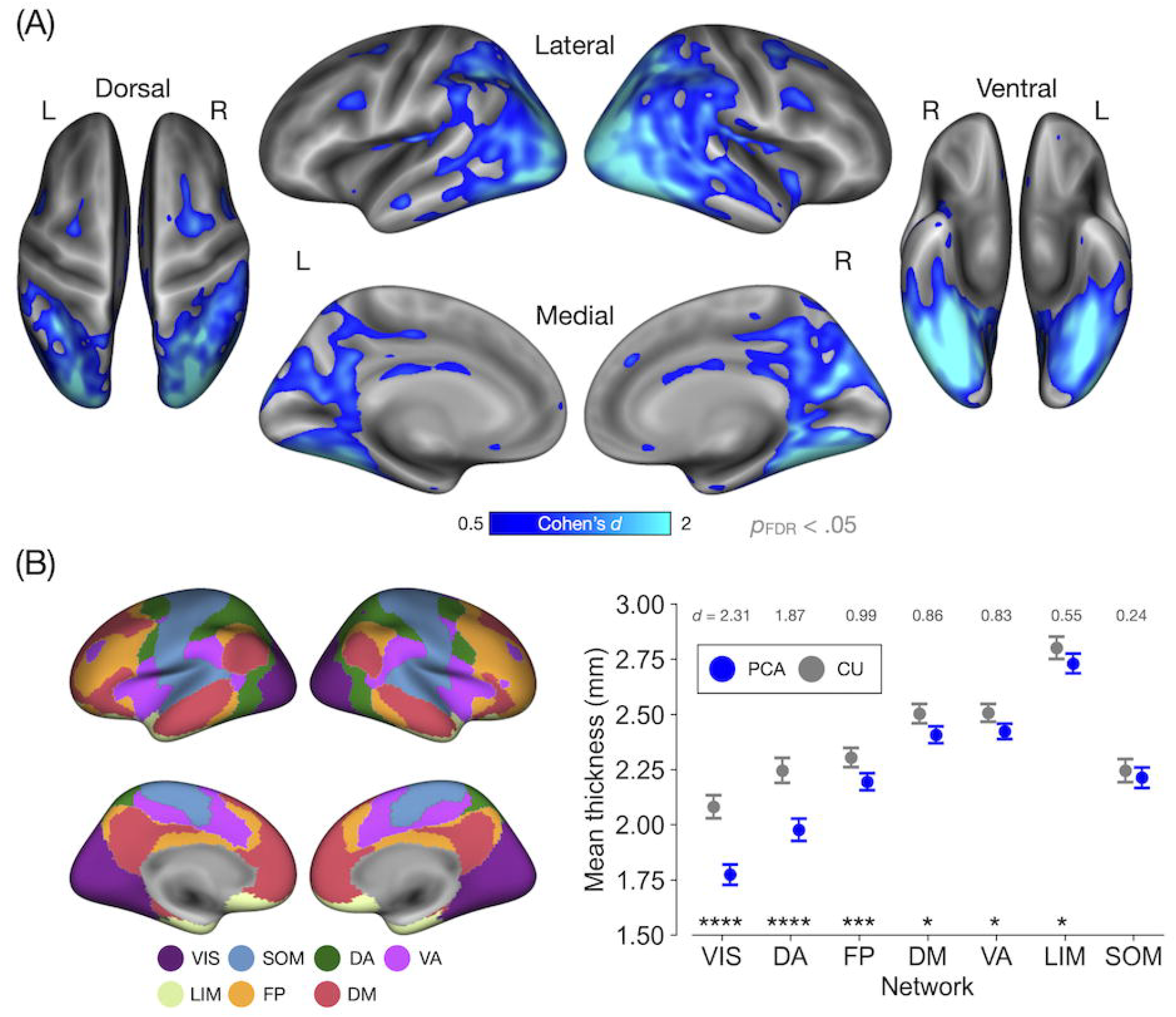
Spatial topography of baseline cortical atrophy in PCA. (A) Colored vertices on the cortical surface indicate areas where PCA patients (*n* = 34) showed cortical atrophy compared with Aβ-CU participants (*n* = 24). Statistical significance for group differences was assessed using a vertex-wise threshold corrected for multiple comparisons by controlling for false discovery rate (FDR) at *p* < .05. (B) Colored vertices on the cortical surface represents a parcellation into seven canonical functional networks^30^. These network labels were used to compute mean cortical thickness within each network, depicted in point plots to the right as estimated marginal means in PCA and Aβ-CU participants. Error bars denote upper and lower bounds of 95% confidence intervals. Gray text shown at the top denotes Cohen’ *d* effect size for each group comparison. VIS = visual; DA = dorsal attention; FP = frontoparietal; DM = default mode; VA = ventral attention; LIM = limbic; SOM = somatomotor. \*\*\*\**p* < .0001, \*\*\**p* < .001, \**p* < .05.

### Baseline cortical atrophy in the DAN predicts longitudinal clinical decline

Vertex-wise linear mixed-effects models identified cortical regions where the magnitude of atrophy at baseline predicted the rate of subsequent change in CDR-SB scores in PCA patients. Compared with the spatial topography of baseline atrophy depicted in **Figure 2A**, we found lesser involvement of the occipital cortex and greater involvement of medial and lateral parietal and lateral temporal areas (**Figure 3A**) in predicting longitudinal clinical decline. At the level of functional networks, we found that the DAN was the only network where the magnitude of baseline atrophy predicted the rate of longitudinal change in CDR-SB scores, as revealed by a significant Time*Thickness interaction: *F*(1,32.49) = 9.06, *p* < .005. Examination of estimated marginal means of linear trends revealed that PCA patients with more prominent DAN atrophy at baseline (as defined by 1 standard deviation [*SD*] below the mean) show a greater increase in CDR-SB scores (i.e., faster clinical decline) by 1.23 points per year compared with those with minimal atrophy in this network (as defined by 1 *SD* above the mean) (**Figure 3B**). No significant Time*Thickness interaction was found in any of the other networks: Visual (*F*(1,32.67) = 0.40, *p ≤* .53), somatomotor (*F*(1,32.66) = 0.90, *p ≤* .35), ventral attention (*F*(1,35.61) = 1.44, *p ≤* .24), limbic (*F*(1,31.15) = 1.94, *p ≤* .17), frontoparietal (*F*(1,39.36) = 0.62, *p ≤* .44), default mode (*F*(1,36.30) = 0.61, *p ≤* .44).

**Figure 3.**
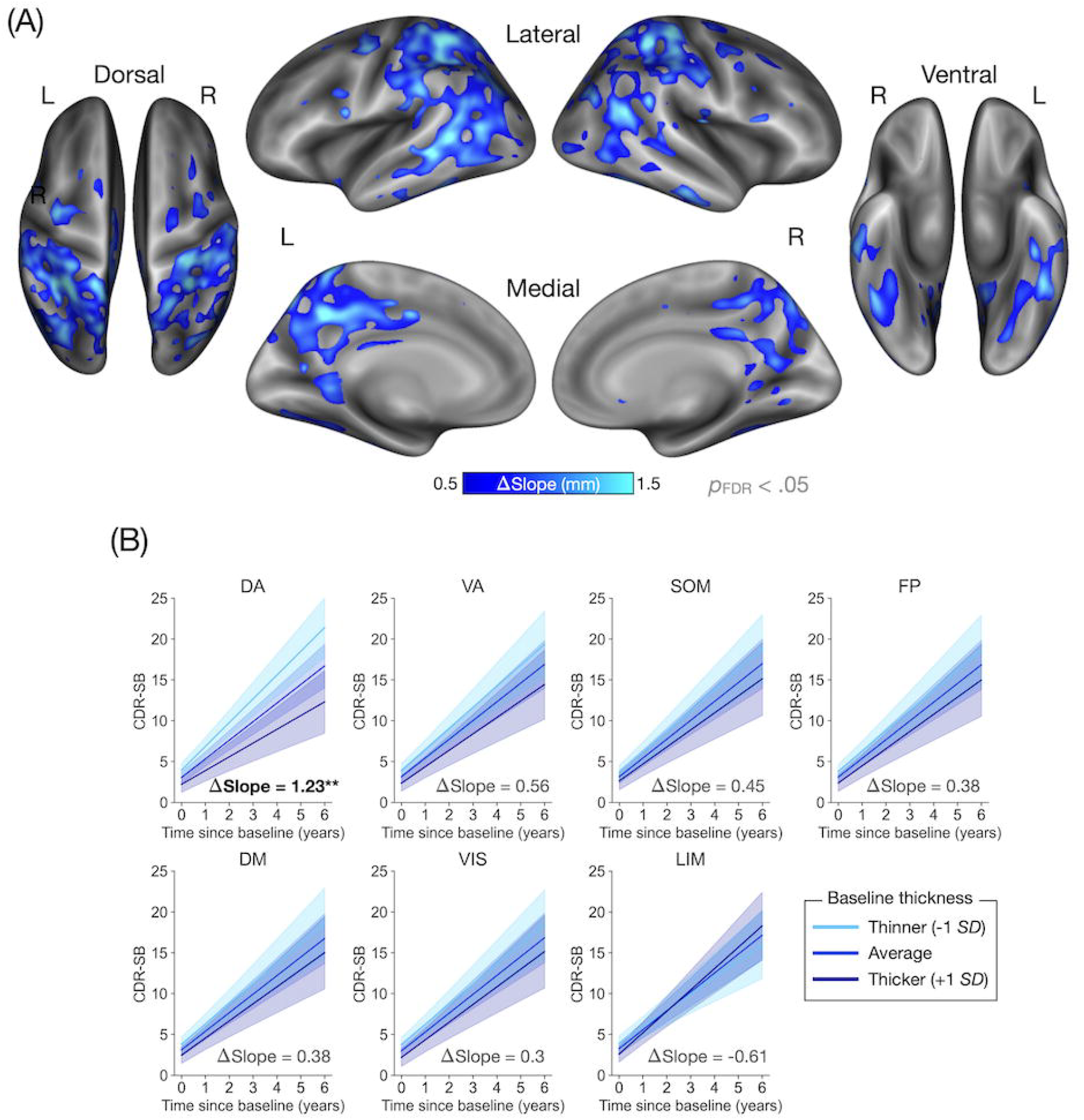
Baseline cortical atrophy predicts longitudinal clinical decline in PCA. (A) Colored vertices on the cortical surface indicate areas where greater atrophy (i.e., decreased thickness) at baseline predicted faster clinical decline as measured by change in CDR-SB scores in our PCA patients (*n* = 34). Vertex-wise values represent the difference in the estimated rates of change in CDR-SB scores (ΔSlope) between one standard deviation (*SD*) above and below the mean of baseline cortical thickness in the population. Statistical significance was assessed using a vertex-wise threshold corrected for multiple comparisons by controlling for the FDR at *p* < .05. (B) Line plots show predicted cortical thickness at each timepoint for each of the seven canonical cortical functional networks, separately for levels of baseline cortical thickness at one *SD* above and below the mean. Shaded bands denote 95% confidence intervals. VIS = visual; DA = dorsal attention; FP = frontoparietal; DM = default mode; VA = ventral attention; LIM = limbic; SOM = somatomotor. \*\**p* < .005.

### Comparison of baseline atrophy vs. atrophy predicting clinical progression in PCA

Finally, we performed an additional comparison of vertex-wise linear regression and mixed-effects model results reported above by projecting the associated statistical maps on a flat map representation of the cerebral cortex (**Figure 4**). These maps further illustrate a clear pattern suggesting anterior progression in the spatial distribution of atrophy in PCA. Specifically, while baseline cortical atrophy was most prominent in posterior cortical regions largely corresponding to the visual network, relatively faster clinical decline was more strongly predicted by atrophy in more anterior regions including the DAN and the neighboring areas within the parietal and temporal lobes, including nodes of the frontoparietal, default mode, and somatomotor networks. At the whole network level, the magnitude of baseline cortical atrophy was greater in the visual network (*M*_*Z*_ = 1.11 ± 1.09) than in the DAN (*M*_*Z*_ = 0.73 ± 0.78) (*p*_perm_ < .0001). In contrast, the magnitude of the relationship between baseline cortical atrophy and the rate of longitudinal change in CDR-SB scores was stronger in the DAN (*M*_*Z*_ = 0.86 ± 0.69) than in the visual network (*M*_*Z*_ = -0.06 ± 0.81) (*p*_perm_ < .0001). Taken together, these findings lend support to the hypothesis that PCA patients whose cortical atrophy has already spread beyond the visual network are more likely to experience faster cognitive and functional decline. As a supplementary analysis, we constructed and evaluated all statistical models based on a subset of the sample with biomarker or pathological evidence consistent with AD pathology (*n* = 29); this analysis yielded results very similar to those reported here in the main text (see Supplemental Material).

**Figure 4.**
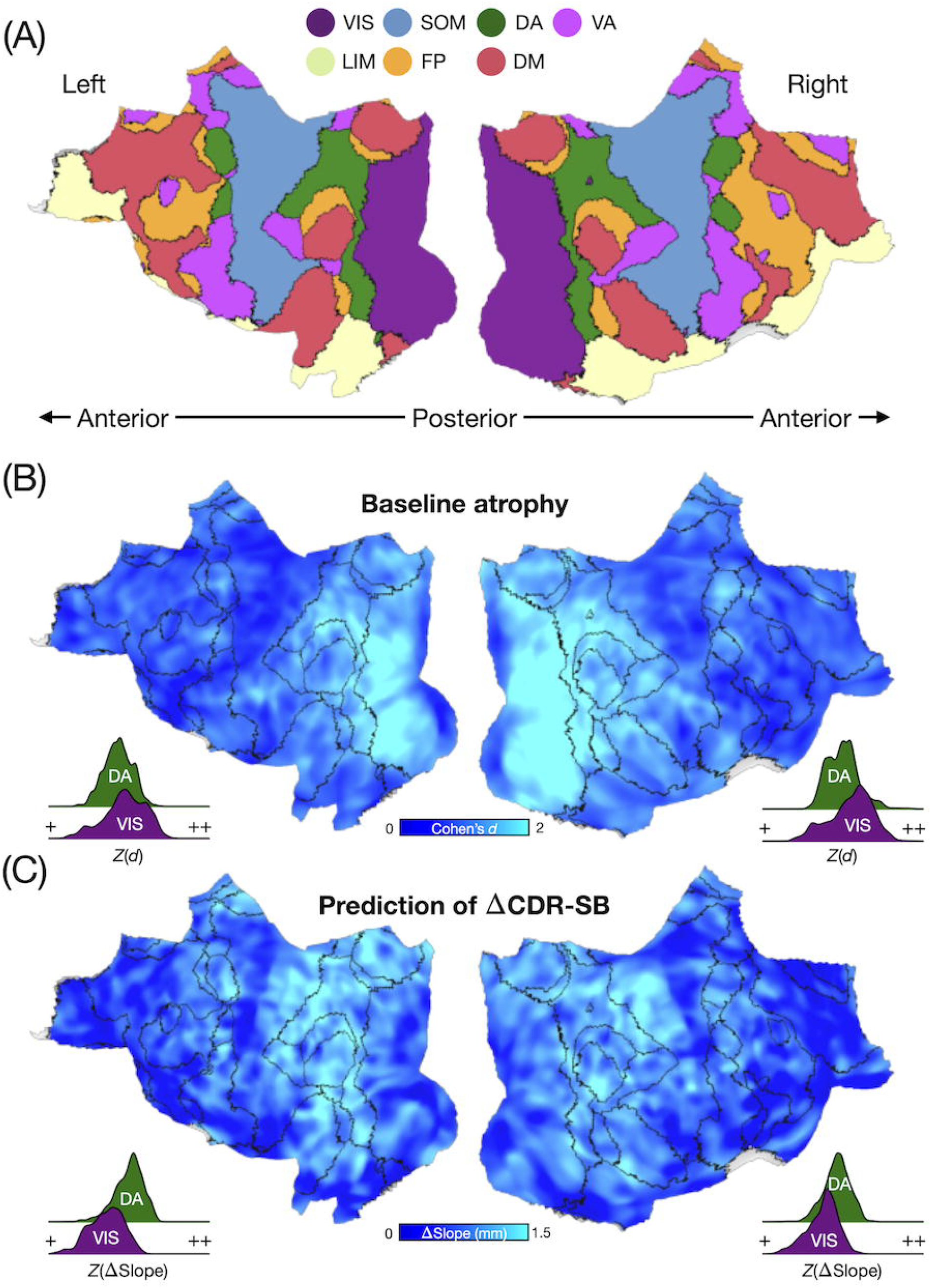
Network-based characterization of baseline cortical atrophy and its utility for predicting clinical decline in PCA. (A) This map illustrates the boundaries of the seven canonical networks^30^ on two flat maps representing the two cerebral cortical hemispheres; see **Figure 2B** for the conventional cortical surface representation of these networks. The maps in (B) and (C) show unthresholded vertex-wise maps of effect size estimates. The first is a Cohen’s *d* map representing the magnitude of difference in baseline cortical thickness between PCA patients and Aβ-CU participants (B). The second is a ΔSlope map representing the difference in the rates of longitudinal change in CDR-SB scores between greater and lesser baseline atrophy relative to the mean (C). A joy plot (ridgeline plot) in each panel in (B) and (C) represents the distribution of vertex-wise effect size values within the DAN (green) and visual network (purple), revealing the more prominent involvement of visual network atrophy at baseline compared with the more prominent involvement of DAN atrophy in the prediction of longitudinal clinical decline. A more rightward distribution indicates overall larger effect sizes.

## Discussion

The hallmark of PCA is predominant visuospatial and visuoperceptual impairment as the earliest signs of illness^3,24^. As PCA progresses, these visual symptoms inevitably worsen and progression of the disease causes impairment in other domains such as memory, language, and executive functions, thus leading to loss of independent function. While some suggest that many PCA patients have a protracted clinical course extending well over a decade^36^, it remains unclear how variable the rate of clinical decline could be across individual patients and what predicts the rate of this decline. In this study, we examined the utility of baseline cortical atrophy for predicting future clinical decline in a sample of individuals with PCA in relatively early clinical stages (CDR 0.5 or 1.0). We found that cortical atrophy localized to the DAN selectively predicted the rate of subsequent clinical decline as measured by changes in CDR-SB scores over clinical follow-up spanning an average of ∼3 years. We propose that this MRI-based biomarker could be used as a valuable clinical prognostication tool that facilitates personalized care planning.

To the best of our knowledge, only one study has reported on trajectories of longitudinal clinical decline as measured by the CDR-SB specifically among individuals with PCA. In a sample of PCA patients (*N* = 28), Whitwell et al.^8^ estimated the annualized rate of change in CDR-SB scores to be 1.15, which is smaller than our estimate (2.28). PCA patients in this study (median baseline CDR-SB = 3.25; Q1, Q3 = [2.5, 5]) and those in Whitwell et al.^8^ (median baseline CDR-SB = 3.00; Q1, Q3 = [2.0, 4.5]) showed comparable levels of clinical impairment at baseline, leaving it unlikely that baseline CDR-SB scores explain this discrepancy. One notable difference between the two studies concerns the number of timepoints from which the annualized rate of CDR-SB change was estimated. In our study, we analyzed CDR-SB scores obtained over ∼3 years on average, with some patients followed over ∼6 years, whereas each PCA patient in Whitwell et al.^8^ was assessed only at two timepoints ∼1 year apart. The rate of clinical progression measured from two timepoints may be noisy^37^; it is thus possible that longer follow-up enabled more stable estimation of the rate of change in CDR-SB scores in our sample. The magnitude of clinical decline we identified was also slightly greater than that observed in samples of older individuals with amnestic AD^38,39^, consistent with evidence that some phenotypes of early-onset AD (including PCA) follow a more aggressive clinical course^40,41^.

Our results indicated that PCA patients with more severe clinical impairment at baseline tended to show faster clinical decline over time. Although this effect was not statistically significant at a conventional threshold, it is consistent with available evidence based on clinical progression in PCA^8^ as well as more heterogeneous samples of individuals with atypical AD including PCA^10,15^. These findings collectively support the utility of baseline cognitive and/or functional assessments for predicting future decline in the same measures. It is noteworthy that some of these studies of atypical AD including PCA have identified the effect of baseline imaging measures on predicting subsequent decline while statistically controlling for baseline clinical assessments, thus highlighting their complementary contributions to clinical prognostication^10,15^. Our data suggest considerable heterogeneity in the rate of clinical decline across individual patients with PCA, even though they were all in the relatively early stages of the clinical syndrome at baseline. The magnitude of baseline cortical atrophy in specific functional networks of the brain may thus be useful in estimating person-specific trajectories of clinical decline as the disease progresses, providing more support for the notion of syndromic heterogeneity within the PCA diagnosis.

The observed spatial topography of baseline cortical atrophy in PCA patients relative to Aβ-CU participants is consistent with prior work identifying predominant involvement of posterior cortical regions at the earliest symptomatic stages of disease^5,17,42–44^. In the frontal cortex, we found modest atrophy selectively localized to bilateral frontal eye fields and inferior frontal junction. The frontal eye fields are considered a key anterior node of the DAN^45^, with the inferior frontal junction also commonly exhibiting intrinsic functional connectivity and task-related co-activation with the DAN nodes^30,46,47^. Although much of the prior research on PCA emphasizes the involvement of posterior cortical regions, there is evidence of hypometabolism^48^, hypoperfusion^49^, and atrophy^50^ in the frontal eye fields. Additionally, molecular imaging studies have demonstrated that tau pathology spreads anteriorly to the frontal cortex over time^19,20^. There is converging evidence that neurodegeneration follows this same anterior progression; relatively greater rates of longitudinal gray matter atrophy have been reported in the temporo-parietal cortices, medial parietal cortices, and frontal eye fields in PCA patients over time compared with the occipital regions where neurodegeneration is typically first observed to occur^5,19,20^. It is therefore likely that some of the PCA patients in our sample were at the early stages of this anterior progression of neurodegeneration at baseline.

Our findings emphasize the importance of the DAN in PCA clinical progression. Although we found suprathreshold vertices within the boundaries of multiple networks (involving the default mode, frontoparietal, and somatomotor networks), this effect mainly emerged as a double dissociation between the involvement of the visual network and the DAN: The greatest magnitude of cortical atrophy was found in the visual network at baseline, though it was atrophy in the DAN that was the primary predictor of longitudinal clinical decline. These results suggest that the visual network in the occipital cortex undergoes neurodegeneration early in the disease course, leaving little variability across individual participants in the magnitude of atrophy at baseline. In contrast, the involvement of the DAN early in the course of PCA is more variable and is reflected in the severity of visuospatial cognitive impairment across individual participants. We recently showed that a cross-sectional index of posterior-to-anterior tau PET signal within the DAN was associated with the magnitude of visuospatial attention deficits in PCA^18^. The current results extend our understanding of the importance of the DAN in PCA by showing the utility of DAN atrophy as a neuroanatomical predictor of the emergence of future cognitive and functional impairment.

We recently published a parallel finding in patients with PPA^14^. In a sample of PPA patients at the MCI stage of symptoms, the language network was most atrophic at baseline compared with control participants. In predicting the rate of progression to dementia, baseline atrophy in the frontoparietal network emerged as a more powerful measure than atrophy in the language network; these results are in line with our prior work on amnestic MCI^9,11^. Taken together, these findings consistently provide support for the notion that the functional brain network(s) reflecting the core syndromic features of a neurodegenerative dementia phenotype may not be the best candidates for prognostication. Instead, patients at an early stage of syndromic cognitive impairment who are beginning to show evidence of neurodegeneration that has progressed beyond the core brain network are at greatest risk for faster cognitive and functional decline in the near future.

Our study has some limitations that warrant acknowledgment and may offer possible avenues for future research. First, our analysis of MRI data was restricted to those obtained at baseline. Longitudinal neuroimaging data—including MRI and PET measures—would be useful in more comprehensively characterizing dynamic trajectories of AD neuropathologic changes and neurodegeneration that may offer additional information beyond a single snapshot for predicting individual clinical trajectories. Second, while the CDR-SB is an established tool widely used as a clinical outcome measure in therapeutic trials and observational studies, this measure was designed for the typical amnestic syndrome of AD. Thus, it does not capture the types of cognitive and functional impairment prominently observed in early stages of PCA (i.e., there is no visuospatial box score). We are currently in the process of refining and validating a specific clinical instrument to fill this need—the Visuospatial Impairment Rating (VIR) scale^17^— that we hope will function as an adjunct to complement the traditional CDR and be sensitive to early dysfunction in PCA for use as outcome measures in imaging-based clinical prognostication. Finally, PCA is a relatively rare clinical syndrome of AD and thus the sample size in this study was also modest. Future work with larger samples, facilitated by data pooling from multiple sites^2^, would allow for out-of-sample testing, assessments of model robustness, and analyses of clinical phenotypes to better investigate PCA heterogeneity.

In sum, these findings provide additional support for the utility of atrophy measures from baseline MRI in prognostication for patients with early-stage cognitive impairment due to AD or other related dementias (ADRD) that will eventually progress. The field now needs to better develop the infrastructure and expertise to bring these kinds of measures into clinical practice, since MRI scans are routinely collected clinically in patients with characteristics like those of the present study. The use of disease signature^9,11^ or functional network-based measures^14^ of brain structure likely have distinct applications for diagnosis, prognostication, or outcomes monitoring given a growing body of evidence supporting its clinical use as we move toward precision medicine in AD and ADRD.

## Data Availability

Data collected and analyzed in this work may be shared (anonymized) at the request of any qualified investigator for purposes of replicating procedures and results.

## Acknowledgments

The authors would like to thank the patients and families who participated in this research, without whose partnership this research would not have been possible. This research was supported by NIH grants R01 DC014296, R01 NS131395, R01 AG081249, K01 AG084820, R21 AG080588, K23 AG065450, K23 DC016912, P01 AG005134, and P30 AG062421 and by the Tommy Rickles Chair in Primary Progressive Aphasia Research. This research was carried out in part at the Athinoula A. Martinos Center for Biomedical Imaging at the MGH, using resources provided by the Center for Functional Neuroimaging Technologies, P41 EB015896, a P41 Biotechnology Resource Grant supported by the National Institute of Biomedical Imaging and Bioengineering (NIBIB), National Institutes of Health. This work also involved the use of instrumentation supported by the NIH Shared Instrumentation Grant Program and/or High-End Instrumentation Grant Program; specifically, grant number(s) S10 RR021110, S10 RR023043, S10 RR023401.

