## Supplemental Material for "Greater baseline cortical atrophy in the dorsal attention network predicts faster clinical decline in Posterior Cortical Atrophy"

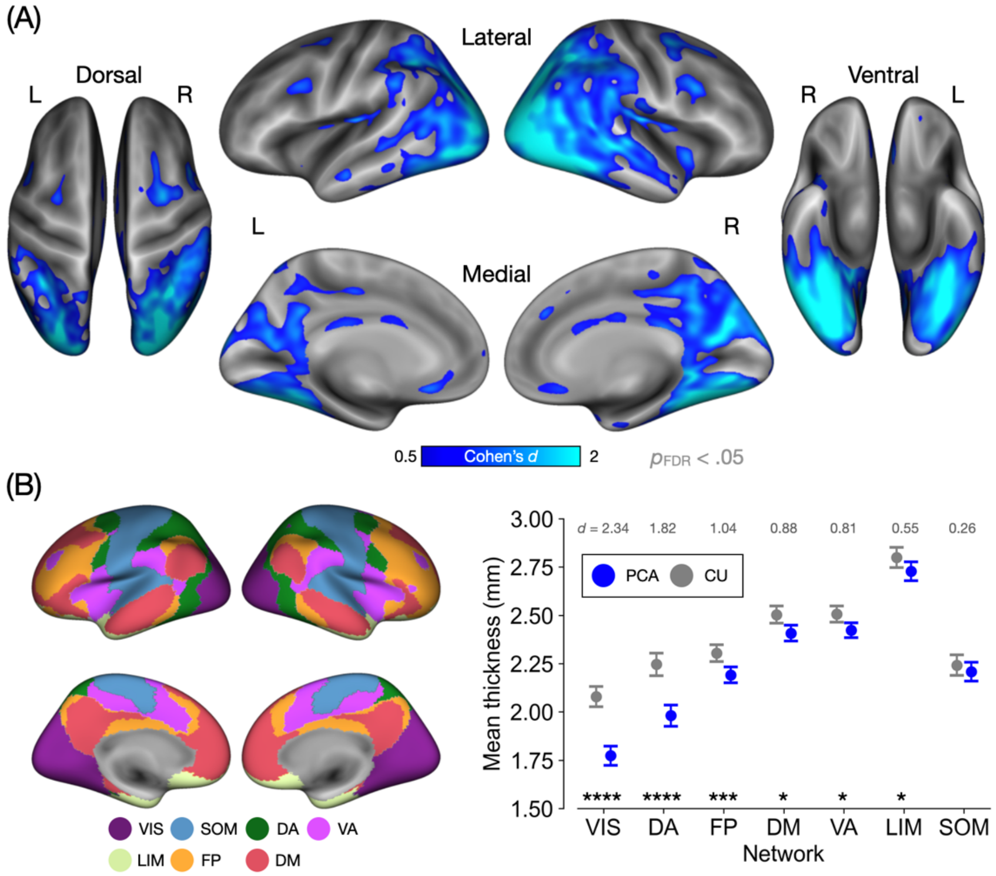


**Supplemental Figure 1. Spatial topography of baseline cortical atrophy in PCA (*n* = 29).** See **Figure 2** in the main text for a detailed description of the underlying analysis. Here, the analysis was based on a subset of PCA patients (*n* = 29) with biomarker or pathological evidence consistent with AD pathology.

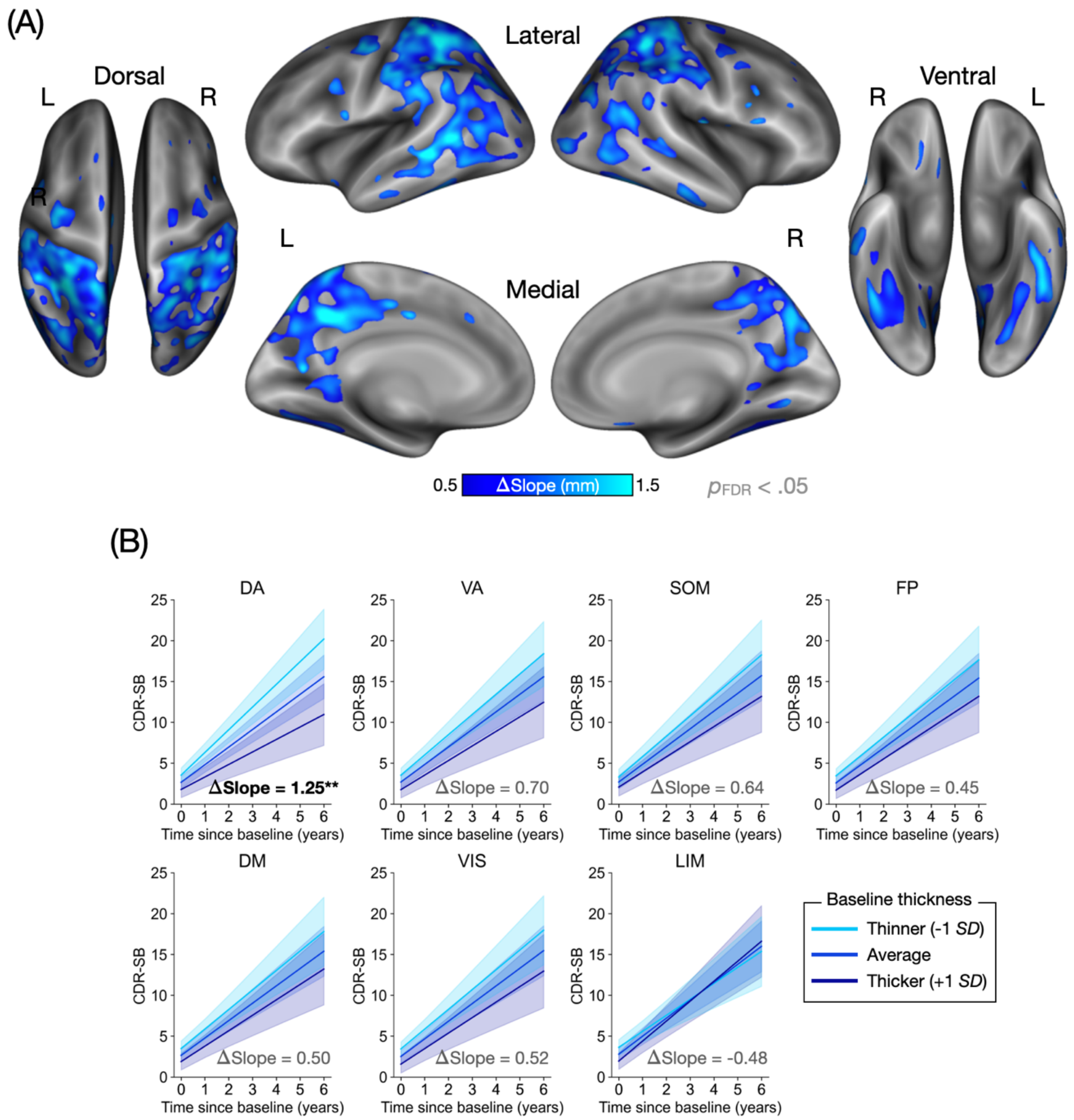


**Supplemental Figure 2. Baseline cortical atrophy predicts longitudinal clinical decline in PCA (*n* = 29).** See **Figure 3** in the main text for a detailed description of the underlying analysis. Here, the analysis was based on a subset of PCA patients (*n* = 29) with biomarker or pathological evidence consistent with AD pathology.

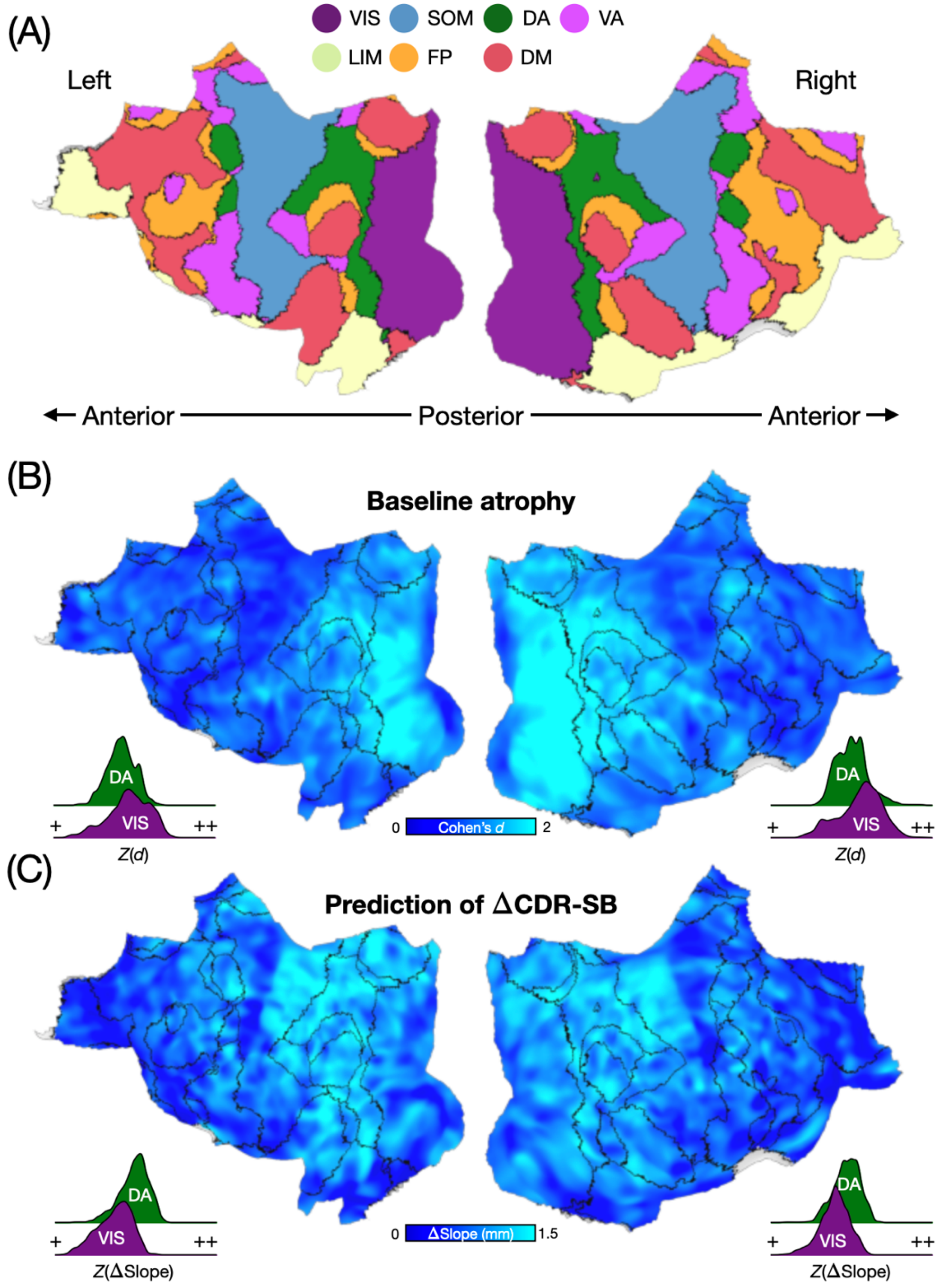


**Supplemental Figure 3. Network-based characterization of baseline cortical atrophy and its utility for predicting clinical decline in PCA (*n* = 29).** See **Figure 4** in the main text for a detailed description of the underlying analysis. Here, the analysis was based on a subset of PCA patients (*n* = 29) with biomarker or pathological evidence consistent with AD pathology.
